# Temperature, humidity, and wind speed are associated with lower Covid-19 incidence

**DOI:** 10.1101/2020.03.27.20045658

**Authors:** Nazrul Islam, Sharmin Shabnam, A Mesut Erzurumluoglu

## Abstract

In absence of empirical research data, there has been considerable speculative hypothesis on the relationship between climatic factors (such as temperature and humidity) and the incidence of Covid-19. This study analyzed the data from 310 regions across 116 countries that reported confirmed cases of Covid-19 by March 12, 2020, and found that temperature, humidity, and wind speed were inversely associated with the incidence rate of Covid-19 after adjusting for the regional and temporal trend in the incidence of Covid-19, columnar density of ozone, precipitation probability, sea-level air-pressure, and length of daytime.

## Background

Despite a considerable amount of interest and hypothesis, empirical evidence on the association between Covid-19 and the meteorological factors (e.g., temperature, humidity) is lacking,^1^ many of which has been reported to be associated with chronic and infectious diseases including severe acute respiratory syndrome (SARS), and Middle East respiratory syndrome (MERS).^2–5^

## Objective

In this study, we examined the association between (concurrent and historical) meteorological factors and the incidence of Covid-19 in 310 regions from 116 countries with reported cases of Covid-19 by March 12, 2020.

## Methods

Reported number of Covid-19 cases were drawn from the database developed by the Johns Hopkins University (data available at https://github.com/CSSEGISandData/COVID-19). We collated the concurrent and historical data since January 08, 2020 on temperature, relative humidity, wind speed, and ultraviolet (UV) index (data available at https://github.com/imantsm/COVID-19) – our primary meteorological factors of interest. We used multilevel mixed effects negative binomial regression model^6^ to examine the association between these factors and the incidence of Covid-19 with a two-level random intercepts nested within the regions (countries, or states/provinces for larger countries). We also conducted two additional analyses to examine the association between historical weather (preceding 1- and 2-week to coincide with the 2019-nCoV incubation period)^7^ and Covid-19 incidence. All the models were adjusted for the regional and temporal trend and variability in the Covid-19 incidence, and other meteorological factors (such as columnar density of total atmospheric ozone, precipitation probability, sea-level air pressure, and daytime length). The effects estimates are reported as adjusted incidence rate ratio (IRR) and their corresponding 95% confidence interval (CI). Data analysis was conducted in Stata/SE 14.2 (StataCorp. 2015. College Station, TX: StataCorp LP).

## Results

Overall, the incidence of Covid-19 increased by 11% (adjusted IRR: 1.11, 95% CI: 1.10-11, P<0.001) per day after adjusting for all the variables listed above (dew point and cloud cover were initially considered, but later excluded due to multicollinearity). In the adjusted model, daily maximum temperature, relative humidity, and wind speed were associated with a lower incidence of Covid-19 (Table 1). There was an inverse association between Covid-19 incidence and 14-day lagged UV index (but not with the concurrent or the 7-day lagged data).

**Table 1.** Association between meteorological factors and incidence of Covid-19.

| Variable | Concurrent |  | 7-day lagged |  | 14-day lagged |  |
| --- | --- | --- | --- | --- | --- | --- |
|  | IRR (95% CI) | P-value | IRR (95% CI) | P-value | IRR (95% CI) | P-value |
| Daily maximum temperature* | 0.94<br>(0.90-0.99) | 0.018 | 0.83<br>(0.78-0.87) | <0.001 | 0.87<br>(0.82-0.92) | <0.001 |
| Relative humidity** | 0.97<br>(0.93-1.00) | 0.052 | 0.97<br>(0.93-1.00) | 0.057 | 0.92<br>(0.89-0.96) | <0.001 |
| Wind speed (m/s) | 0.96<br>(0.94-0.98) | 0.001 | 0.93<br>(0.91-0.96) | <0.001 | 0.93<br>(0.90-0.95) | <0.001 |
| UV index | 0.98<br>(0.96-1.01) | 0.268 | 1.01<br>(0.98-1.04) | 0.510 | 0.94<br>(0.91-0.97) | <0.001 |

-The models were additionally adjusted for the temporal trend in the incidence of Covid-19, columnar density of ozone, precipitation probability, sea-level air-pressure, and length of daytime; IRR: Adjusted incidence rate ratio; CI: confidence interval
\* Estimated effects with every 5°C increase in temperature; \*\* Estimated effects with every 10% increase in humidity

## Discussion

To our knowledge, this is the first empirical study to comprehensively examine the association between the incidence of Covid-19 and a range of meteorological factors spanning across the world. Our finding of an inverse relationship between temperature (and humidity) and the incidence of Covid-19 may suggest a cold and dry environment more favourable condition for virus survival, as was proposed for other coronavirus such as SARS-CoV and MERS-CoV.^2–5^ An inverse association with wind speed may indicate a shorter suspending time in the air due to dilution and removal of Covid-19.^3^ An inverse association with a higher UV index would suggest viral destruction at higher temperature,^3^ but the association did not hold with the concurrent or 7-day UV index.

## Data Availability

Available on request

